# Magnetic resonance imaging of the gastrointestinal tract shows reduced small bowel motility and altered chyme in cystic fibrosis compared to controls

**DOI:** 10.1101/2021.02.15.21251749

**Authors:** Neele S Dellschaft, Christabella Ng, Caroline Hoad, Luca Marciani, Robin Spiller, Iain Stewart, Alex Menys, Helen Barr, Penny A Gowland, Giles Major, Alan R Smyth

## Abstract

Cystic fibrosis (CF) is a genetic disease affecting mucosal secretions. Most patients experience digestive symptoms, but the mechanisms are incompletely understood. Here we explore causes and consequences of slower gastrointestinal transit using magnetic resonance imaging (MRI).

Twelve people with CF and 12 healthy controls, matched for age and gender, underwent MRI scans both fasted and after standardised meals over a period of 6.5 hours. Images were assessed for small bowel motility, longitudinal relaxation time (T_1_) of ascending colon chyme, chyme texture and appearance of the colon wall.

Small bowel motility scores were significantly lower in CF than in healthy controls in the fasting state (CF median 40 arbitrary units IQR [31, 46] vs Control 86 a.u. [52, 106], *P*=0.034). This difference was less pronounced postprandially. Furthermore, ascending colon chyme T_1_ was lower in CF than in controls (CF 0.59 s [0.38, 0.77] vs Control 0.79 s [0.55, 1.31], *P*=0.010). The difference in texture between small bowel and colon chyme, seen in health, was diminished in CF (difference in Haralick contrast 0.90 a.u. [0.38, 1.67] vs Control 2.11 a.u. [0.71, 3.30], *P*=0.010). Ascending colon mucosa in CF participants had an abnormal appearance compared to controls (Score 1-3, CF 2 [1, 3] vs Controls 1 [1, 1], *P*=0.019).

Reduced small bowel motility and water content of ascending colon chyme are consistent with slower transit and constipation. MRI provides unique insights into chyme texture in the small bowel (suggesting bacterial overgrowth) and the appearance of the colon mucosa (suggesting altered mucus) in CF.

**Key point summary:**

- People with cystic fibrosis (CF) have intrusive digestive symptoms and severe gut complications, but mechanisms are incompletely understood.
- In this study, 12 people with CF were compared to healthy controls, undergoing repeated MRI scans before and after standardised meals.
- Fasted small bowel motility is reduced in people with CF, consistent with slower transit. In addition, a reduced colonic chyme water content and abnormal appearance of small bowel and colonic chyme as well as colonic mucosa suggest small bowel bacterial overgrowth, fat malabsorption and abnormal mucus production
- These MRI outcomes hold promise in the assessment of therapeutic interventions.

## Introduction

Cystic fibrosis (CF) is the most common, life-limiting, autosomal, recessive disorder in people of northern European ancestry. The genetic mutation causing a defect in the cystic fibrosis transmembrane conductance regulator protein (CFTR) drastically reduces chloride transport function (Elborn, 2016). This reduces secretions, making mucus more viscous. Since lung infections are the main cause of morbidity and mortality in people with CF, the respiratory system has received the majority of research and treatment attention. As a result, people with CF now live longer and, the median life expectancy for a baby born with CF in the UK today is 49 years (Cystic Fibrosis Trust, 2020). With life expectancy increasing, more attention is now directed to improving morbidity.

Gastrointestinal symptoms present a high burden for people with CF (Tabori *et al*., 2017; Hayee *et al*., 2019). The mechanisms behind these symptoms are poorly understood and there are limited treatment options (Rowbotham *et al*., 2018), in part because our understanding of the pathophysiology is incomplete. Malabsorption is treated by giving pancreatic enzymes in capsules (pancreatic enzyme replacement therapy). A high fat diet, combined with higher doses of pancreatic enzyme replacement therapy, in people with CF has been associated with increased life expectancy of around ten years (Corey *et al*., 1988). Nevertheless, people with CF still commonly experience symptoms such as constipation, diarrhoea, abdominal pain, bloating, nausea and excessive flatus (Smith *et al*., 2020).

CF distinctly affects gastrointestinal physiology: In nearly all people with CF there is a failure of secretion of pancreatic enzymes and bicarbonate into the small bowel (Cystic Fibrosis Genotype-Phenotype Consortium, 1993). This means the neutralisation of gastric acid no longer occurs as normal in the proximal duodenum. Food and especially fat will be poorly digested, causing energy deficiency and impaired growth (Chase *et al*., 1979). Bile acid secretion (Weizman *et al*., 1986) as well as intestinal re-absorption (Weber *et al*., 1976) are disturbed. Intestinal inflammation is common, especially in patients with pancreatic insufficiency (Bruzzese *et al*., 2014).

The most severe complication (occurring in 6% of patients every year (Cystic Fibrosis Trust, 2020)) is a partial or complete blockage of the bowel by a build-up of sticky material in the terminal ileum – distal intestinal obstruction syndrome (DIOS).

Computed tomography (CT) imaging of gas bubbles in the dilated, obstructed small bowel of people with CF suggests the presence of colonic bacteria and faecal material (Mayo-Smith *et al*., 1995). There is general clinical consensus that small bowel bacterial overgrowth is commonly present in people with CF, but it is difficult to obtain objective evidence (Dorsey & Gonska, 2017) as conventional glucose breath hydrogen tests are insensitive and relate poorly to symptoms (Lisowska *et al*., 2009).

The Gut Imaging for Function and Transit in Cystic Fibrosis Study aimed to gain insights into how gastrointestinal physiology differs in people with CF compared to unaffected controls using serial magnetic resonance imaging (MRI) scans, both fasted and in response to meals. Our group has extensive experience investigating gastrointestinal function in health and disease using MRI (Spiller & Marciani, 2019). Common scan sequences range from describing anatomy, organ volumes and chyme water content (Major *et al*., 2018) to cine sequences allowing assessment of intestinal motility (Khalaf *et al*., 2019).

We have previously reported primary and secondary outcomes from this study (Ng *et al*., 2020), in which a higher small bowel water content suggested impaired stimulation of ileal emptying after eating (the gastro-ileal reflex (Deiteren *et al*., 2010)) and obstructed transfer of small bowel chyme to the colon.

Here, we report exploratory outcomes testing the hypotheses that small bowel motility is reduced in CF which may result in prolonged small bowel transit (Ng *et al*., 2020) compared to healthy controls, and that there is a difference in small bowel and colon chyme compared to that of healthy controls. Chyme water content as well as the composition of the chyme are considered, using subjective scoring and objective quantitative methods.

## Methods

This study was carried out as described previously (Ng *et al*., 2020). In short, 12 people with CF (homozygous for p.Phe508del mutation, with a forced expiratory volume in 1 second (FEV_1_) over 40% of predicted, 12-40 years of age; see (Ng *et al*., 2020) for complete list of criteria and Table 1 for demographics) and 12 age and gender matched healthy controls participated in this study. None of the CF participants had CF-related diabetes, and only one had a history of DIOS. All people with CF were recruited through Nottingham University Hospitals NHS Trust and controls were recruited by advertisement in Nottingham, UK. Participants were asked not to exercise on the day before their study scans, and not to take their normal laxatives and anti-diarrhoeals. All scans took place at the Sir Peter Mansfield Imaging Centre, University of Nottingham, and scans were acquired on a 3T Philips Ingenia scanner (Philips Healthcare, Best, the Netherlands) using a torso coil.

**Table 1:** Demographics of study participants.

| **Median [IQR]** | **CF (n=12)** | **Control (n=12)** | **P-value** |
| --- | --- | --- | --- |
| **Age (years)** | 19 [15, 24] | 19 [15, 25] | 0.42 |
| **Male, n (%)** | 7 (58) | 7 (58) |  |
| **Weight (kg)** | 56 [53, 59] | 62 [59, 75] | 0.005 |
| **Height (cm)** | 166 [158, 175] | 166 [160, 176] | 0.33 |

Participants attended in the morning after an overnight fast. They were given two standardised meals during the day, as used in previous studies (Marciani *et al*., 2010; Chaddock *et al*., 2014; Major *et al*., 2018; Wilkinson-Smith *et al*., 2019): Meal 1 consisted of 300 g creamed rice pudding, 25 g seedless raspberry jam, 30 g double cream, with a drink of 100 mL orange juice and 240 mL water (all Sainsbury’s®, UK; 519 kcal, fat 19 g, carbohydrate 77 g); Meal 2 was given 270 minutes later and consisted of 400 g macaroni cheese (Sainsbury’s®, UK;), 100 g strawberry cheesecake (Rhokett®, UK) and 240 mL water (total 1110 kcal, fat 54 g, carbohydrate 116 g;). CF participants took pancreatic enzyme replacement therapy with meals as prescribed.

### Measurements

#### Blinding

All images were assigned a code number before analysis, blinding researchers to disease status.

#### Motility

Small bowel motility was assessed on 6 slices (2 coronal slices acquired over one minute with a temporal resolution of one second, using a free breathing cine balanced turbo field echo (bTFE) sequence) at time points t −30 (fasted), 0, 30, 60 and 300 minutes from the first meal (see Fig 1). Only the time points closest to the meals, t −30, t0 and t300 were assessed for differences between CF and Controls.

**Figure 1:**
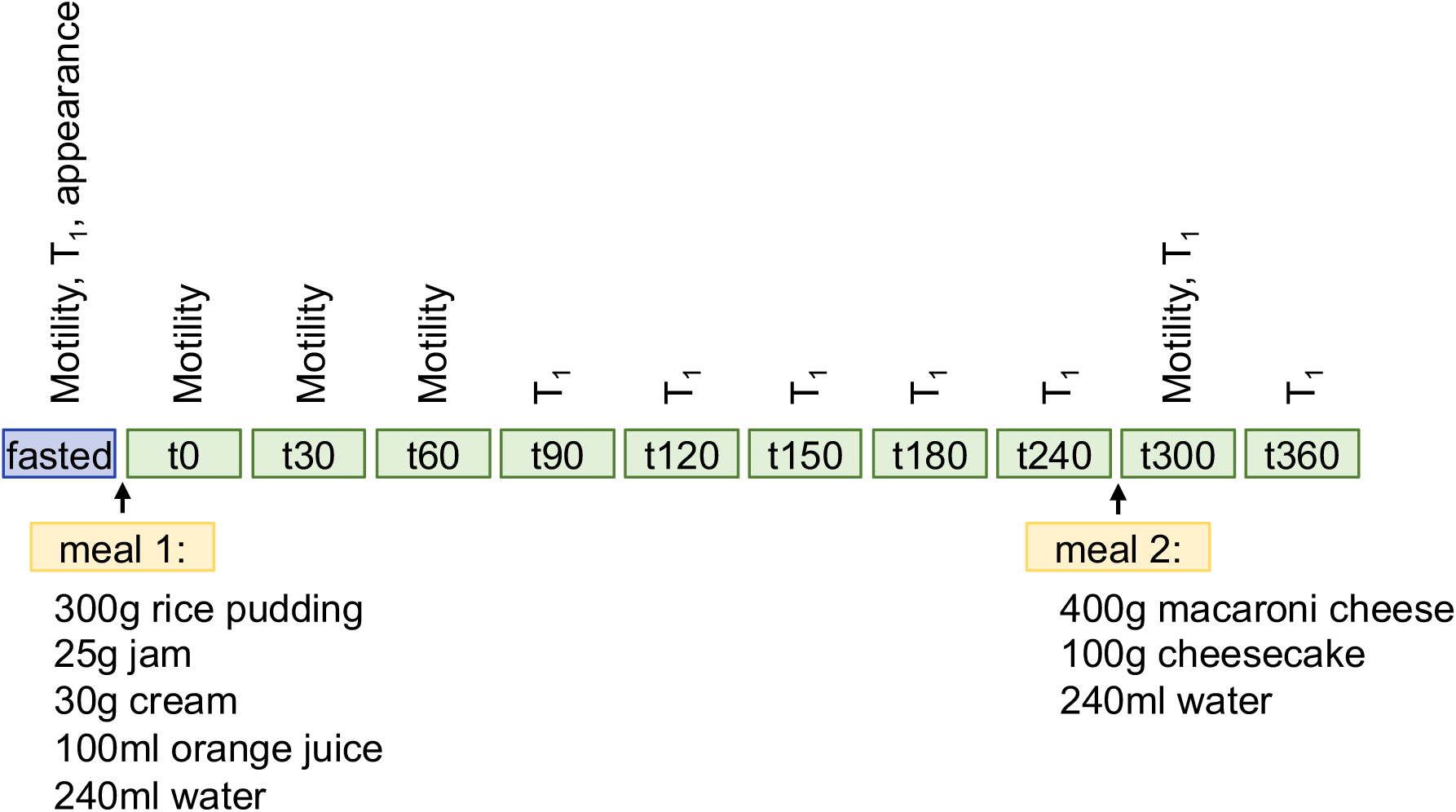
Diagram of study day procedures. Participants arrived fasted and were scanned right away. They then received meal 1 (300 g rice pudding, 25 g jam, 30 g cream, 100 ml orange juice, 240 ml water), followed by scan t0, then half-hourly scans until 180 minutes after the meal (t180), followed by three more hourly scans, with meal 2 (400 g macaroni and cheese, 100 g cheesecake, 240 ml water) given after t240. Study participation was complete after scan t360. Scans were taken for motility, T_1_ and analysis of appearance at the time points indicated.

Motility was assessed as previously described (Menys *et al*., 2014; Khalaf *et al*., 2019). Cine MR images were adjusted for respiratory motion using GIQuant® (Motilent, London, UK). Regions of interest (ROI) of the small bowel were drawn on each slice, and movement within these regions was tracked over time (nonlinear optic flow registration). Changes in signal intensity (in mathematical terms, power spectrum of registration parameter C, representing a summary of through-plane motion and flow of chyme from bowel wall movement (Menys *et al*., 2014)) were measured and further analysed using a customised graphical user interface written in MATLAB (MathWorks, Natick, MA, USA).

#### T_1_ of colonic chyme

Longitudinal relaxation rate, T_1_, is the decay time constant for the MRI signal. It is a measure of the time taken for spinning protons to realign with the external magnetic field after excitation. T_1_ of the chyme in the ascending colon was measured using a single slice bTFE sequence with preparatory 180 degree inversion pulse applied before acquiring the imaging data (Marciani *et al*., 2014); data were acquired from 8 different inversion times ranging 100-5000 ms at time points t-30 (fasting) and t90-t360. Both motility scans and T_1_ scans are quite time-consuming so we prioritised motility scans in the early postprandial phase when motility is most stimulated, and T_1_ in the later phase, seeing that T_1_ is relatively stable over time. Only the time points closest to the meals, t - 30, t240 and t300 were assessed for differences between CF and Controls.

T_1_ was measured in three small ROIs per participant and time point, avoiding areas with obvious gas, and then averaged for each time point. By determining the mean signal from each ROI at the 8 inversion times, data were fitted to a model of the signal evolution to determine the value of T_1_ (data with an R^2^ of less than 0.9 to the model was rejected due to poor fitting). A longer T_1_ indicates a higher water content in the chyme (Major *et al*., 2018), with T_1_ of the liver typically being 800 ms, and in blood typically being 1600 ms (Obmann *et al*., 2020).

#### Texture of bowel contents

Texture of bowel contents was assessed on a dual fast field gradient echo sequence with echo times 1.15 ms (out of phase, T_1_-weighted) and 2.30 ms (in phase), repetition time 110 ms and 60° flip angle. 22 coronal slices (7 mm thick with 1 mm gap) were acquired with voxel size 1.6 × 1.6 × 8 mm at fasting baseline, t-30.

We observed that in CF texture of contents of the small bowel and colon were similar whereas small bowel and colon contents differed in texture in healthy controls. Therefore, we developed a system to score these observed differences subjectively.

Small bowel and colon T_1_-weighted images were scored by three independent assessors (NSD, CH, CN) on a three-point ordinal scale (see Fig 2 for example images). For small bowel, the texture of luminal contents was scored 1 (normal, homogeneous throughout the small bowel), 2 (moderately heterogeneous) or 3 (inhomogeneous, which could include differences in signal brightness or obvious mottling). For colon, the texture of luminal contents was similarly scored 1 (smooth appearance), 2 (moderately heterogeneous) or 3 (normal, mottled appearance). The final score was the score assigned by 2 or more assessors.

**Figure 2:**
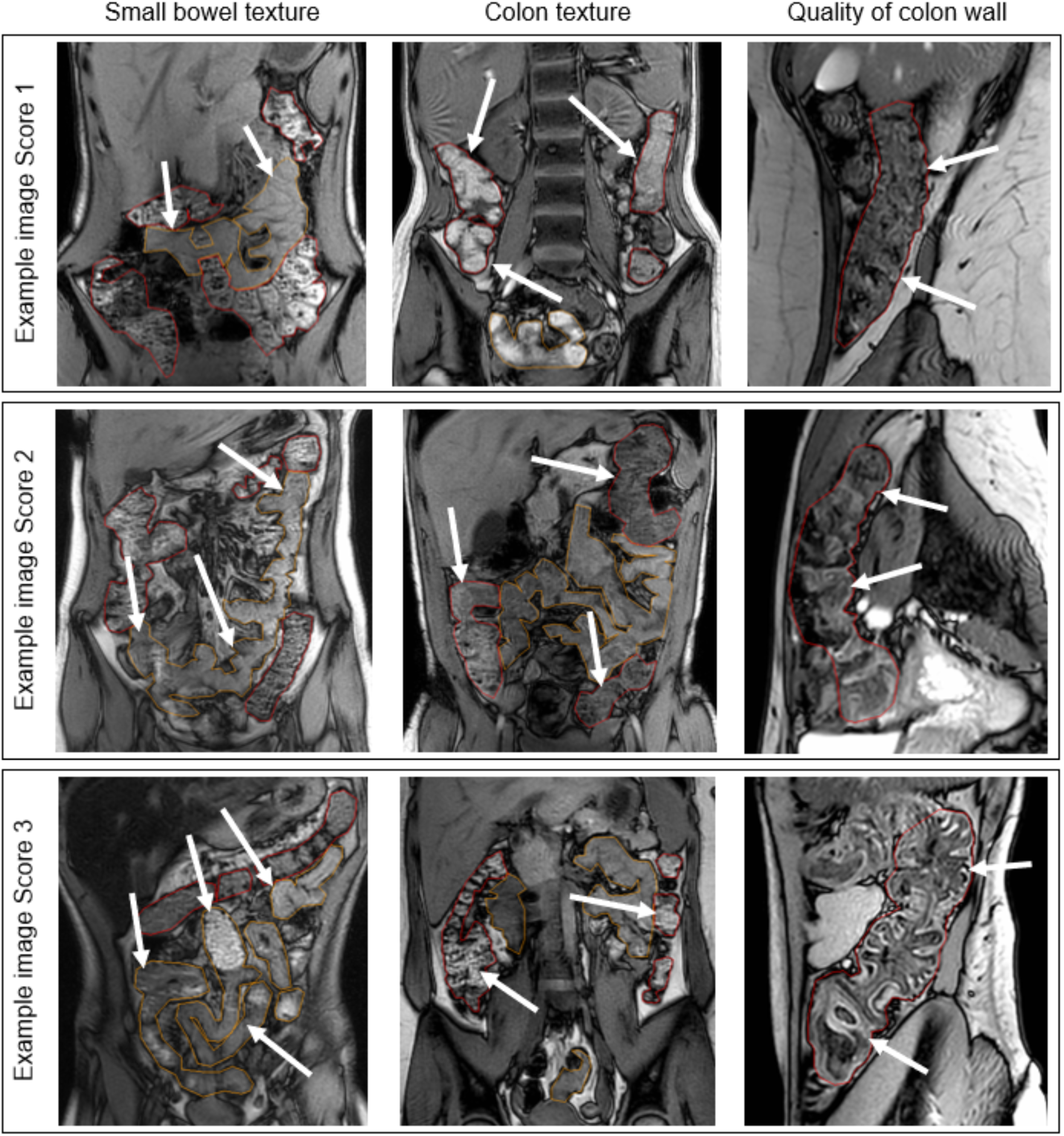
Representative images for semi-quantitative, ordinal scoring of small bowel and colon luminal contents texture as well as sagittal images for scoring of the colonic wall appearance (both healthy and CF images are shown). Left column: Each participant’s small bowel content was visually scored between 1 (normal, homogeneous appearance) and 3 (very heterogeneous). Middle column: Colonic texture was scored between 1 (smooth appearance) and 3 (normal, mottled appearance). Right column: Each participant’s ascending colon was assessed for appearance of bowel wall, with scores between 1 (normal appearance: colon wall as thin dark line) and 3 (colon wall shows as thick white line with clear haustral folds). White arrows point to the tissues of interest, i.e., small bowel in the left column, and to colon in the middle and right columns.

Texture of small bowel and colonic luminal contents was also assessed quantitatively on the same images to provide a more objective measure of the changes. Haralick contrast maps (Haralick, 1979; Hall-Beyer, 2017) were calculated in MIPAV (Center for Information Technology, US National Institutes of Health, Bethesda, MD, USA) from the T_1_-weighted images in two dimensions, spatially invariant, with an offset of 1 (i.e., voxels compared are immediate neighbours in all directions), window of 7×7 voxels, 32 grey levels. Higher values correspond to regions with mottled texture with great contrast between adjacent pixels, lower values to smooth texture (see Fig 3). ROIs were drawn for the whole small bowel as well as for the colon, avoiding obvious gas pockets and partial volume effects, and then eroded (10 mm sphere) to exclude any contamination from gut wall.

**Figure 3:**
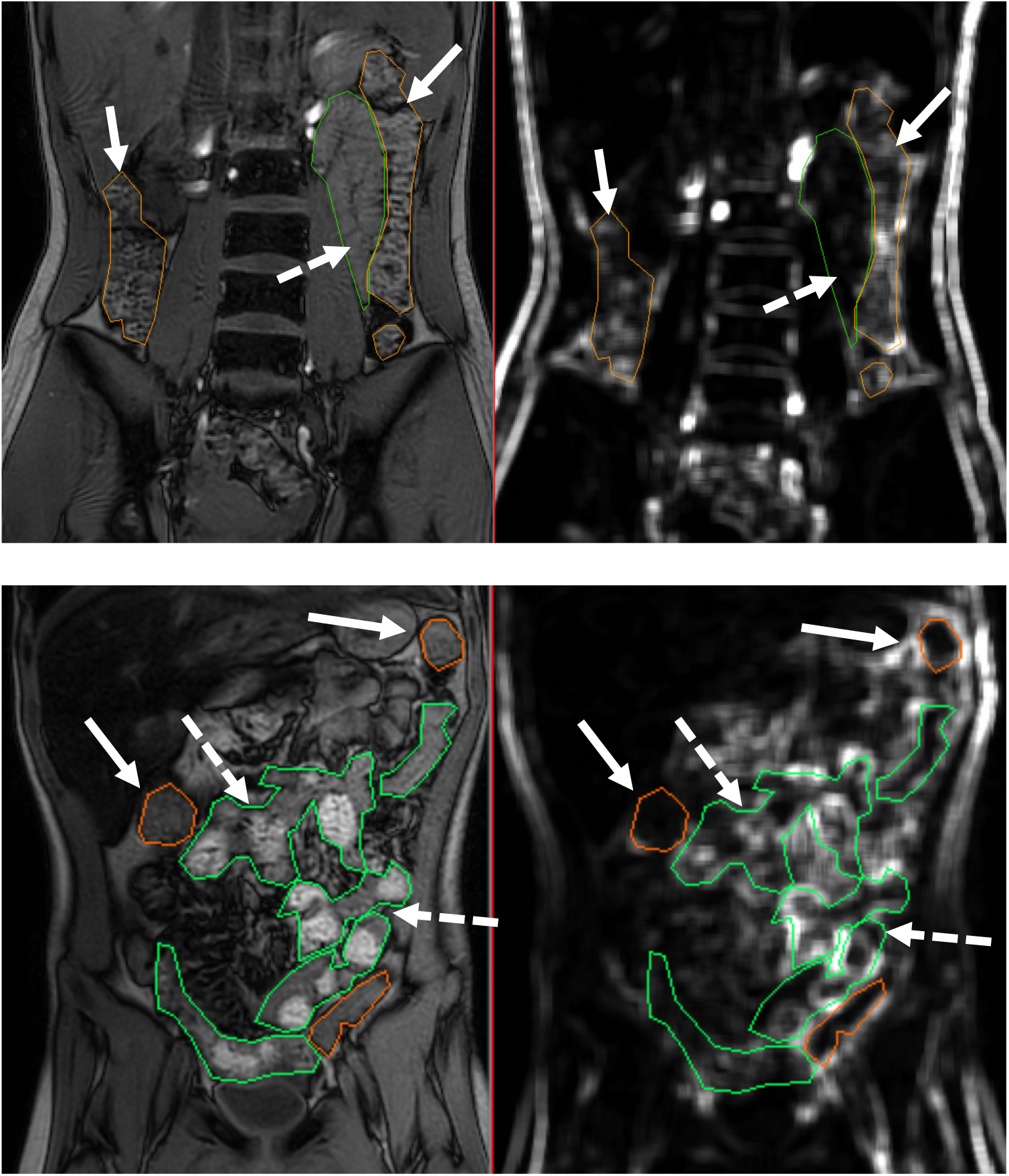
Demonstration of Haralick contrast maps to analyse texture of luminal contents of small bowel and colon. A coronal T_1_-weighted image was calculated into a Haralick contrast map. Example shown in the top row is a healthy control participant with typical textures in both small bowel (dashed arrow) and colon (solid arrows), whereas the example in the bottom row is a participant with CF with heterogeneous, rather mottled texture in the small bowel (dashed arrows) and smooth texture in the colon (solid arrows).

#### Colon mucosa

For observation of ascending colon changes, a high-resolution balanced turbo field echo sequence with echo time 1.2 ms, repetition time 2.4 ms and 42° flip angle, 8 sagittal slices (7 mm thick with 0.6 mm gap) was acquired with voxel size 1.5 × 1.5 × 7 mm at fasting baseline, t-30.

It was observed that a subset of scans had an unusual thick white line on the luminal side of the colon wall. Therefore, we developed a second ordinal scoring system to describe observed changes in the appearance of the colonic wall. As above the images were scored as 1 (normal, colon wall shows as a thin dark line), 2 (some signs of white outline) or 3 (colon wall shows as a thick bright line, following the haustral folds; see Fig 2 for example images). A total of three assessors scored the appearance of the ascending colon wall and the final score was the score assigned by 2 or more assessors.

### Statistics

All continuous data was considered for statistical significance (*P*<0.05) using non-parametric Wilcoxon tests. To minimise multiple comparison, analyses of motility and T1 between CF and age/sex matched controls were restricted to three timepoints of before meal 1 (fasted), after meal 2 (t300), and either after meal 1 (t0, for motility) or before meal 2 (t240, for T_1_).

Ordinal image score distribution between disease statuses were assessed using Fisher’s exact tests. Cronbach’s alpha statistics were used to evaluate internal consistency between assessor scores, alpha >0.7 was considered acceptable agreement. All statistical analysis were performed with SPSS V24 (IBM, Armonk, NY, USA).

This study was approved by the UK National Research Ethics Committee (18/WM/0242) and conformed to the standards set by the Declaration of Helsinki. All participants gave written informed consent or, if a participant was under 16, parents gave informed consent with the child’s assent. The study was registered on ClinicalTrials.gov NCT03566550.

## Results

### Motility

Small bowel motility was significantly lower in people with CF at fasting baseline (*P* = 0.034; Fig 4; please see appendix, Table S1 for all medians, IQRs and *P*-values; see example motility videos in the Appendix). Consuming meal 1 raised motility in both groups but the difference between them was less obvious immediately after the first meal owing to high variability within normal controls (*P* = 0.084). Motility was similar between groups after the second, larger meal (*P* = 0.433). Fig 4 shows examples of motility intensity maps.

**Figure 4:**
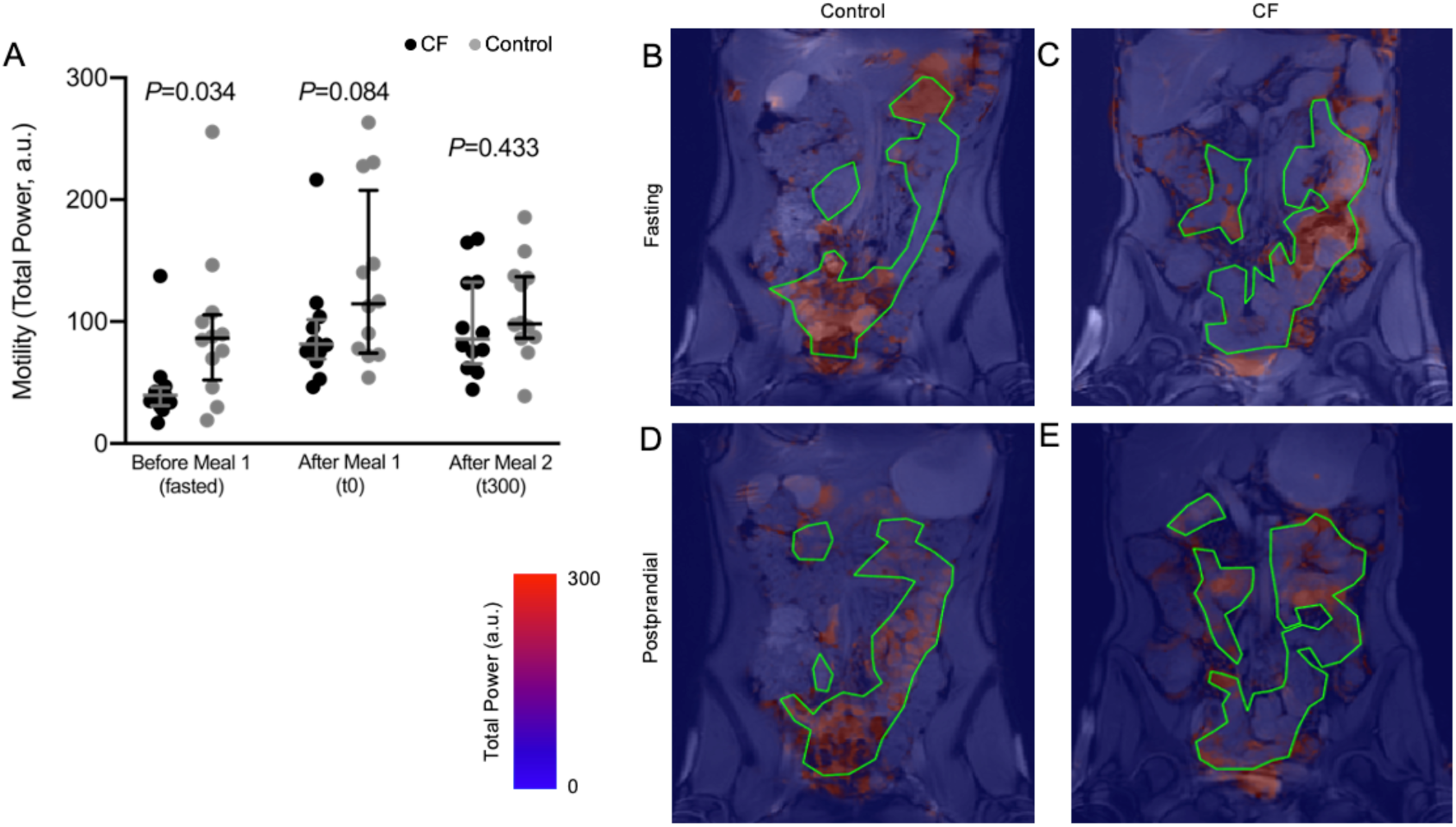
Small bowel motility as assessed on cine MRI. This measure represents the signal intensity changes from both bowel wall movement and chyme bolus movement through the small bowel. Values are median and IQR as well as all data points. B) and C) are example motility maps at baseline, D) and E) at t0, with scale bar given. B) and D) are a healthy control participant and C) and E) a participant with cystic fibrosis. Green outlines indicate small bowel.

### T_1_ of colonic chyme

The longitudinal relaxation time in the ascending colon when fasted was significantly lower in CF (*P* = 0.010) but not before or after the second meal (*P* = 0.182 and *P* = 0.594; Fig 5).

**Figure 5:**
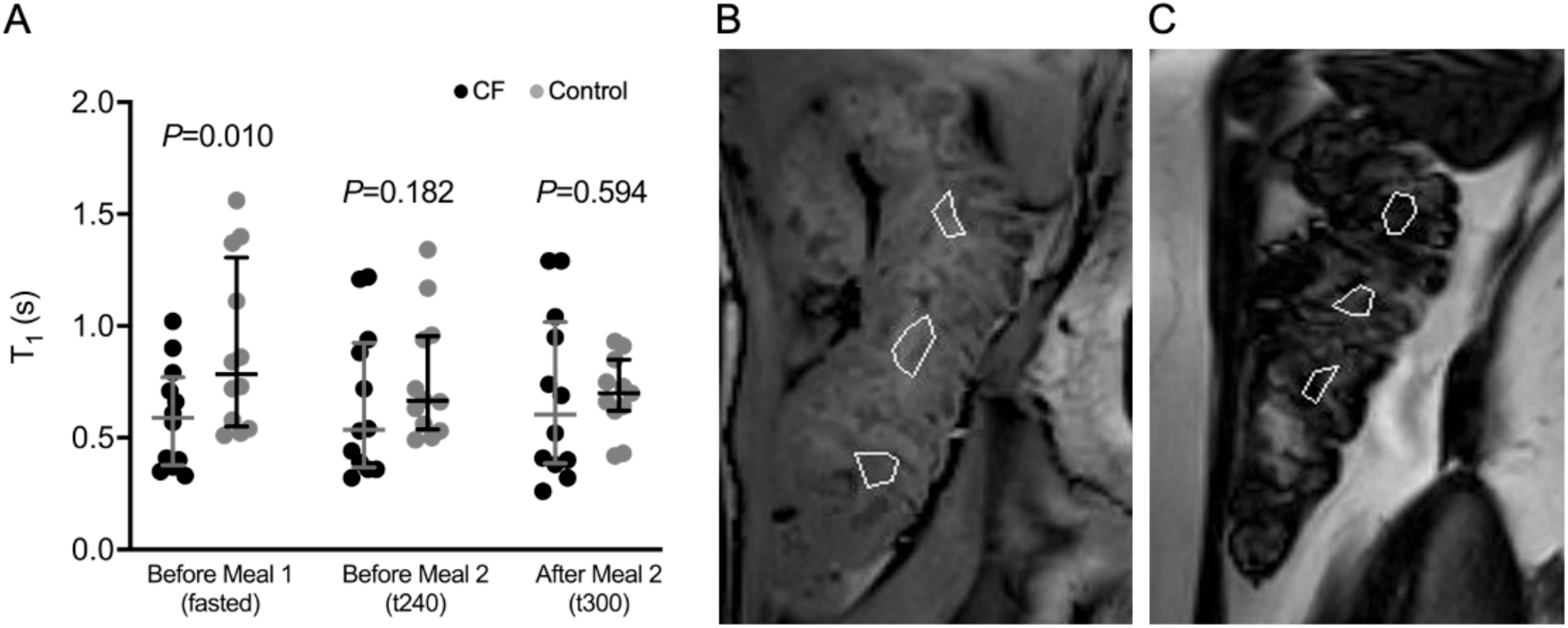
T_1_ of colonic chyme. Values are median and IQR as well as all data points. Example images show placement of ROIs to measure T_1_ in participants B) with CF and C) control participants, data from which are then averaged before analysis.

### Texture of bowel contents

Agreement between the three assessors’ scores was acceptable for both small bowel and colon chyme texture (small bowel chyme texture, Cronbach’s alpha = 0.79; colon chyme texture, Cronbach’s alpha = 0.73).

Assessors’ scoring of small bowel chyme texture was significantly different in people with CF compared to controls (representative images given in Fig 2; Fig 6A and B, Fisher’s exact *P* = 0.013). Colon chyme Haralick contrast was significantly lower in people with CF (*P* = 0.034; Fig 6E). The difference between the Haralick contrasts of chyme in small bowel and colon was significantly less in people with CF than in healthy control participants (*P* = 0.010; Fig 6F) and similar to the equivalent calculated from the ordinal scores (Fig 6C). Semi-quantitative, ordinal score and Haralick score were positively correlated in the small bowel and in the colon (small bowel, Spearman’s rho 0.622, *P*=0.001; colon, Spearman’s rho 0.762, *P*<0.001).

**Figure 6:**
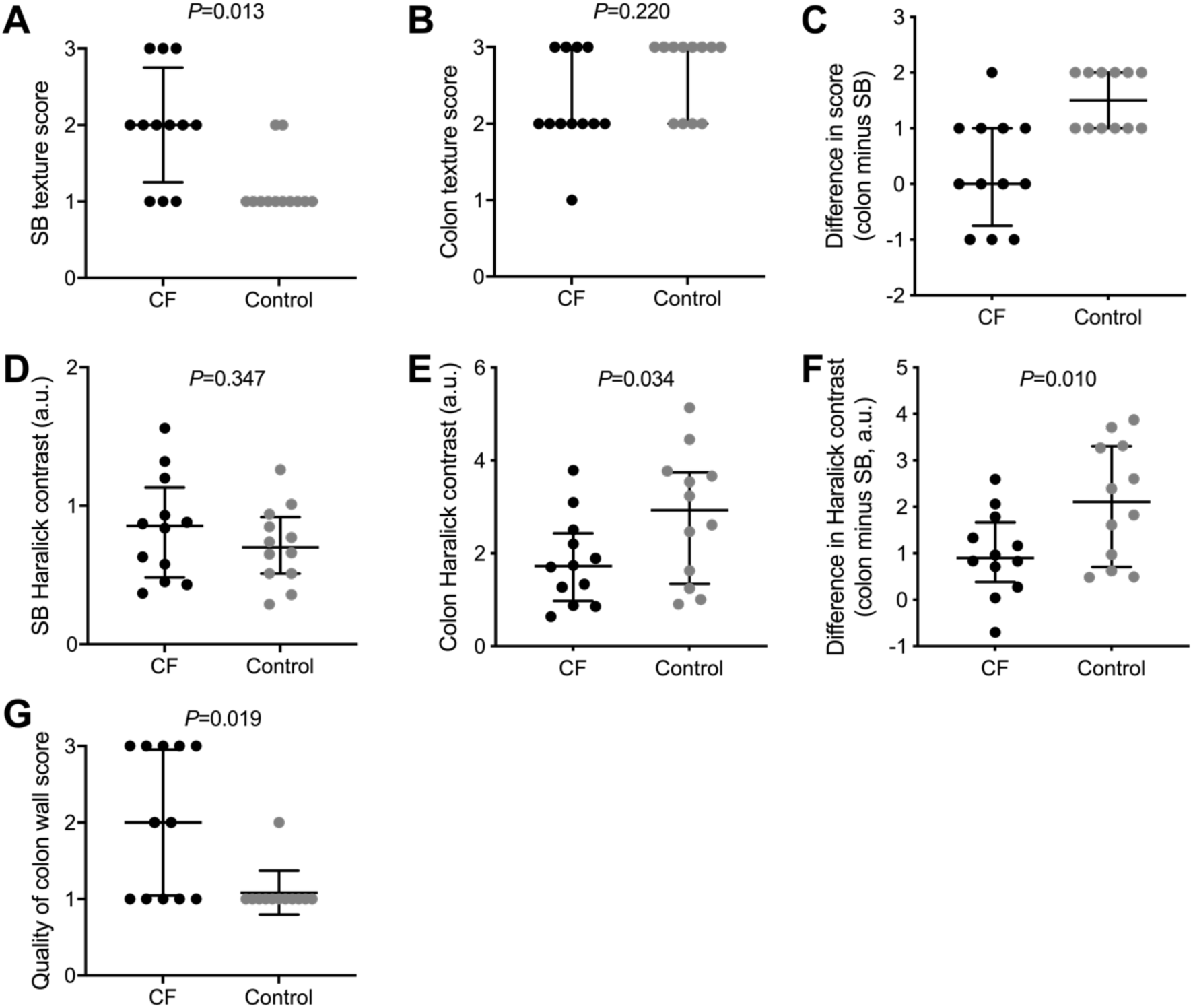
Assessment of small bowel and colon texture and differences between intestinal compartments semi-quantitatively (A-C; no *P*-value given for C as scores are ordinal, serving as a comparison to F) and by Haralick contrast (D-F). Semi-quantitative, ordinal scores for appearance of colon wall (G). In all graphs, median and IQR as well as data points are indicated. *P*-values given are for Fisher’s Exact or Wilcoxon tests.

### Colon mucosa

The appearance of colon mucosa in CF was significantly different from controls (Fig 6G, Fisher’s exact *P* = 0.019) as assessed by semi-quantitative, ordinal scores (good agreement between scorers, Cronbach’s alpha = 0.87).

## Discussion

This study aimed to investigate potential causes and consequences of prolonged oro-caecal transit in CF, which was previously observed by our group (Ng *et al*., 2020) and others (Gelfond *et al*., 2013). Gastrointestinal symptoms in CF are proposed to derive from a cycle of chyme stasis causing bacterial overgrowth, inflammation and further inhibition of motility. We found reduced small bowel motility, reduced T_1_ of the ascending colon in the fasted state, as well as an altered appearance of small bowel and colon chyme and ascending colon bowel walls.

The measurement of motility undertaken in this study will be affected by both small bowel wall movement and bolus movement. This was lower in people with CF. This may be due to change in chyme consistency (caused by maldigestion) and to partial obstruction at the level of the ileocaecal valve. We cannot ascertain whether this reduction is a primary difference in contractile activity or a secondary effect of chronic distension as demonstrated by the higher small bowel water content (Ng *et al*., 2020), chronic inflammation and fibrosis (Bruzzese *et al*., 2014). Postprandial motility in CF appeared to become more similar to healthy controls. This restoration of motility, following a meal, is not seen in Crohn’s patients (Khalaf *et al*., 2020) undergoing MRI (Khalaf *et al*., 2020) – a phenomenon attributed to fibrosis. This suggests that, in contrast to Crohn’s disease, fibrosis does not occur in CF. This area requires further investigation, including markers of small bowel diameters, gut wall thickening, fibrosis and gut hormone feedback loops (Murphy *et al*., 1992).

Malagelada et al. (Malagelada *et al*., 2020) conducted a study using wireless video capsule endoscopy and reported reduced contractile activity and increased pooling of turbid contents in the small bowel of people with CF. This may be comparable to our observations of reduced bolus propulsion and altered chyme texture, and our previously reported observation (Ng *et al*., 2020) that water accumulates in the small bowel in people with CF. However, in the Malagelada study, intestinal motility could only be recorded postprandially, as optical data can only be captured at the camera’s location and all subjects consumed a 300-kcal liquid meal 45 minutes after camera ingestion. In contrast, our study, using cine MRI of the whole small bowel, showed a lower motility, but not increased small bowel water content, in fasted participants. This difference in motility was less clear after the first meal and, of note, we found no difference in motility after our second, larger meal. This may suggest that an appropriate motility response can be induced by a meal larger in calorie content and size.

Ascending colon T_1_, a measure of the water content of chyme, was lower in people with CF when fasting, with values similar to those seen in people with chronic constipation (Major *et al*., 2018). The increased colon volumes that we reported previously (Ng *et al*., 2020) also suggest constipation (Lam *et al*., 2017) which is a common GI problem in people with CF (Smith *et al*., 2020); the presence of inspissated (i.e. less watery) faeces in people with CF has been well documented (Snyder *et al*., 1964). Importantly, the lower T_1_ shows that the higher water content we observed in the small bowel (Ng *et al*., 2020) does not translate to higher colonic water content. This may be due to slow entry of fluid from the small bowel into the colon, allowing more time for water absorption. Additionally, since CFTR inhibits activity of the sodium transporter ENaC, the defective CFTR seen in CF will lead to increased sodium and, with that, water absorption from the lumen (Mall *et al*., 1999).

Review of study images showed some obvious differences between CF participants and healthy controls, which we formalised with a categorisation analysis for assessing the texture in small bowel and colon chyme. There is no objective quantitative method in the published literature for image analysis of the colon mucosa, for comparison with our semi-quantitative, ordinal scores. In CF (but not in control participants) the image textures of colon and small bowel were similar. This finding was consistent both in the semi-quantitative and in the quantitative analysis (using Haralick contrast maps). These findings suggest an abnormal small bowel chyme composition in CF, similar to the contents of the colon. Abnormal small bowel chyme composition may occur in CF due to bacterial overgrowth, as gas bubbles in the small bowel chyme have been observed on CT scans from patients with small bowel obstruction (Mayo-Smith *et al*., 1995). The differences seen, especially those in the colon (where we would expect to see a mottled texture), may be due to fat malabsorption, leading to a more pasty stool texture, altered microbiota (Burke *et al*., 2017), or indeed the altered mucosal secretions. It is worth noting that all patients took their usual doses of pancreatic enzyme replacement therapy with study meals so fat digestion was optimised.

As mentioned above, the build-up of sticky material in the lumen of the terminal ileum – known as distal intestinal obstruction syndrome (DIOS) – is a severe complication seen in around 6% of people with CF per annum (Cystic Fibrosis Trust, 2020). Snyder et al described paediatric cases of DIOS who required surgery where a faecal mass was found in the caecum, extending into the terminal ileum (Snyder *et al*., 1964). Histological findings included a thick layer of viscous mucus lining the mucosa of the terminal ileum and extending as “streamers” into the lumen. We propose that the thick bright line following the haustral folds seen in over half of our CF participants is indicative of this adherent mucus layer. Our findings suggest that these pathological abnormalities, typical of DIOS, may also occur in CF patients who are experiencing less severe gastrointestinal symptoms such as abdominal pain and bloating.

### Strengths and limitations

This study required participants to spend over six hours in the scanning facility. The protocol therefore placed demands both on the participants and the facility, limiting sample size. It remains to be seen whether these changes can be corroborated in a larger number of people with CF. However, our MRI protocol allowed us to describe a wide range of GI characteristics in a way that is non-invasive and well tolerated in people with CF. Since a considerable number of the people with CF had obvious differences in gastrointestinal appearance, as demonstrated with the ordinal scores, assigning code numbers may not have completely blinded the researchers carrying out the image analysis. However, the bias introduced was limited since most analyses used global ROIs of the small bowel or colon. Our quantitative analysis is not part of current clinical practice but allows us to highlight distinctive features which elucidate gastrointestinal pathology in CF. Our research approach requires trained staff for image analysis, is time-consuming and costly. However, we plan to develop a simplified protocol to answer specific clinical questions, as opposed to the detailed characterisation that we have reported here.

## Conclusions

This study is the first to use serial MRI assessment to explore abnormalities of intestinal function in CF. MRI has allowed us to capture multiple physiological parameters which may underpin the gastrointestinal symptoms experienced by people with CF. We have found quantitative and semi-quantitative differences in gut motility, luminal contents and gut wall characteristics between people with CF and healthy controls. These insights provide a basis to assess the GI effects of therapeutic interventions such as CFTR modulators. They also provide a basis for new and refined hypotheses for the treatment of gut related disorders in CF. Simplified protocols will be needed before MRI can fulfil a role in routine clinical assessment, but these data suggest that MRI is a promising tool for assessment of gut function and transit in CF.

## Data Availability

The data that support the findings of this study are available from the corresponding author upon reasonable request.

## Authors’ contribution

The study was designed by AS, GM, RS, LM, ND, PG & CN. MRI scanning protocol design, data acquisition and analysis were conducted by CN, ND, CH, AM, LM, IS & PG. Statistical analysis was by IS & ND and all authors contributed to the interpretation of study data. The first draft of the manuscript was written by ND. All authors have reviewed and approved the manuscript.

## Declaration of Competing Interest

ND, CH, LM, IS & PG have nothing to disclose. CN and GM report grants and speaker honorarium from Vertex, outside the submitted work.

RS reports grants from Zespri International Ltd and Sanofi-Aventis, as well as lecturing fees from Menarini and Alfawasserman, outside the submitted work.

AM is the CEO of Motilent Limited, a medical imaging analysis company.

AS reports grants from Vertex, as well as speaker honoraria and expenses from Teva and Vertex, outside the submitted work. In addition, AS has a patent issued “Alkyl quinolones as biomarkers of Pseudomonas aeruginosa infection and uses thereof”.

