## Supplemental Table 1 for "Magnetic resonance imaging of the gastrointestinal tract shows reduced small bowel motility and altered chyme in cystic fibrosis compared to controls"

### Overview of data

Table S1: Summarised data from all outcomes. *P*-value for Wilcoxon comparisons (continuous measures) and Fisher’s Exact tests (ordinal scores) between CF and control participants, matched for age and sex. Where *P*-values are not given a statistical assessment was not considered to reduce multiple testing. The difference in ordinal scores between colon and small bowel was used only to illustrate that outcomes were similar to the difference in quantitative Haralick scores.

| Median [IQR] | CF | Control | *P*-value |
| --- | --- | --- | --- |
| Small bowel motility (arbitrary units) | | | |
| Fasted | 40 [31, 46] | 86 [52, 106] | **0.034** |
| t0 | 82 [69, 102] | 115 [74, 208] | 0.084 |
| t30 | 59 [50, 73] | 101 [59, 144] |  |
| t60 | 51 [40, 74] | 67 [58, 120] |  |
| t300 | 86 [65, 132] | 98 [86, 137] | 0.433 |
| T_1_ (seconds) | | | |
| Fasted | 0.59 [0.38, 0.77] | 0.79 [0.55, 1.31] | **0.010** |
| t90 | 0.51 [0.38, 1.08] | 0.70 [0.53, 1.27] |  |
| t120 | 0.52 [0.38, 0.98] | 0.79 [0.55, 1.24] |  |
| t150 | 0.60 [0.41, 1.12] | 0.78 [0.56, 1.17] |  |
| t180 | 0.56 [0.40, 0.93] | 0.71 [0.57, 0.86] |  |
| t240 | 0.54 [0.37, 0.93] | 0.67 [0.54, 0.96] | 0.182 |
| t300 | 0.61 [0.39, 1.02] | 0.70 [0.62, 0.85] | 0.594 |
| t360 | 0.53 [0.34, 0.91] | 0.66 [0.47, 0.93] |  |
| Ordinal scores | | | |
| Small bowel chyme texture | 2 [1.25, 2.75] | 1 [1, 1] | **0.013** |
| Colonic chyme texture | 2 [2, 3] | 3 [2, 3] | 0.220 |
| Difference (colon - SB) | 0 [-0.75, 1] | 1.5 [1, 2] |  |
| Appearance of colon wall | 2 [1, 3] | 1 [1, 1] | **0.019** |
| Haralick contrast (arbitrary units) | | | |
| Small bowel chyme texture | 0.85 [0.48, 1.13] | 0.70 [0.51, 0.92] | 0.347 |
| Colonic chyme texture | 1.73 [0.98, 2.43] | 2.93 [1.34, 3.74] | **0.034** |
| Difference (colon - SB) | 0.90 [0.38, 1.67] | 2.11 [0.71, 3.30] | **0.010** |
